# Protocol for a multi-country investigation of overseas care and cancer survivorship in small islands developing states of the Eastern Caribbean: The CaSIDEC study

**DOI:** 10.1101/2025.08.17.25333877

**Authors:** Aviane Auguste, CaSIDEC study group

## Abstract

**Background:** Small Island Developing States (SIDS) make up worldwide, 65 million people. SIDS have some of the highest rates of cancer mortality burden in the developing world. Disparities in cancer mortality could be attributed to reliance on services overseas which is a common feature across SIDS due to limited resources for comprehensive cancer care on-island. Overseas travel for cancer care, remains largely understudied in SIDS, and epidemiological data on the impact on patient outcomes are lacking. We aim to investigate the association with overseas travel for care and known determinants of patient outcomes (treatment delays, lifestyle and social support) in Caribbean SIDS using mixed-methods.

**Methods:** We will establish a cohort of 900 cancer survivors residing in the islands of Antigua, Dominica, Grenada, Saint Kitts, Saint Lucia and Saint Vincent. Eligible participants will be adult cancer survivors (any cancer site, histology and diagnosis year) and having accessed health services in their island of residence due to cancer. Sites for recruitment will be cancer support groups, public and private hospitals and oncology clinics. Participants will be contacted by a trained interviewer for a face-to-face survey. For every country visited for diagnosis and/or treatment, we will record the services accessed, and motives for the choice of country. In-depth interviews and focus groups will be conducted with cancer survivors and their caregivers to learn about overseas travel and social support systems.

**Discussion:** This is the first investigation on the influence of overseas travel on cancer care in multiple Caribbean SIDS. Our work will produce recommendations to fill performance gaps in the systems for cancer care in the OECS that will improve patients’ lives at multiple phases. It is an important first step to developing data resources for high-quality research in understudied SIDS.

## BACKGROUND

### Cancer burden in small island developing states

Populations that are geographically isolated and resource-constrained (GIRC) face heavier cancer burdens compared to those in more populous and affluent settings [1, 2] due to critical performance gaps in cancer control in these GICRs. In GIRC settings, the proportion of patients presenting with advanced-stage cancer are greater and is driven in part by low health literacy and insufficient opportunities for cancer screening. Only 10.6-11.2% of Caribbean women diagnosed with breast cancer, present at stage I, compared with 48% in the USA [3, 4]. The few numbers of oncologists and pathologists available for patient care in these settings also hinders access to life-saving cancer treatment. Little scholarly work exists on cancer control in GIRCS to address these gaps [1, 5, 6]

Among GICRs, Small Island Developing States (SIDS) are a subset that make up worldwide, 65 million people[7]. SIDS have some of the highest rates of cancer mortality burden in the developing world. Even compared to larger low-and-middle-income countries (LMICs) in Africa, age-standardized mortality rates for cancer are higher in small islands from the Caribbean and Polynesia (87.4 vs. 99.2 and 118.9 per 100 000 inhabitants) [8]. Disparities in cancer mortality could be attributed to heavy reliance on care services overseas which is a common feature across SIDS [9, 10] due to limited resources for comprehensive cancer care on-island [11].

### Overseas travel for cancer care and patient outcomes

Referral for cancer care overseas in SIDS is often perceived as an opportunity to access better quality services compared to those available locally; however, little is known about how the process of overseas referral affects subsequent delays in the care continuum. An overseas referral is a complex process in SIDS, and possible benefits from accessing better quality services may be compromised by time delays. Time delays may occur for a myriad of reasons (system, provider or patient related) such as having to mobilize resources to finance the stay overseas, long decision-making intervals, waiting for visa approval, and other logistics [12].

In addition to treatment delays, survivorship care in this context is an issue. The incidence of cancers is expected to increase in SIDS in the coming decades [13]; however, small island health systems are currently not prepared to satisfy the growing demand in survivorship care [5]. Survivorship care is aimed at preventing cancer recurrence, secondary cancers, and improving quality-of-life through healthy lifestyles [14]. Healthy lifestyles can reduce risk of mortality/disability from cancer and treatment-associated problems such as cardio-metabolic and lung disease, bone loss, eye and hearing changes, and lymphedema [15–20]. Regular physical activity and healthier diets can contribute to better long-term health outcomes among cancer patients [21]. Management of these late treatment effects, follow-up care and health promotion services are under-developed in Eastern Caribbean SIDS [1]. Consequently, long-term outcomes for survivors who travelled for care may be better than those who didn’t due to additional health promotion and resources offered by providers overseas compared to those on-island [1, 22, 23]. Disparities in mortality and long-term disabilities among working survivors deprive the Caribbean of productive workers who contribute to much-needed economic growth [24]. However, Caribbean studies on lifestyle factors among cancer survivors are scarce. Previous studies on lifestyle have focused largely on the period before diagnosis [25] which does not reflect lifestyle later during the disease trajectory [26]. Furthermore, the potential disparities between survivorship care done locally and overseas have not been measured.

Social isolation is another issue that may arise from overseas travel for cancer care. This could be counterproductive, leading to worse health outcomes [27, 28]. Social support is defined as a network of family, friends, neighbours, and community members that is available in times of need to give psychological, physical, and financial help [29]. The link between social support and improved quality of life is well established [30–32]. A previous study on breast cancer patients showed that having social support mediates the choice of coping strategies toward positive reframing, which leads to better emotional well-being [31]. The role of social support in overseas medical travel in the Caribbean is unclear but the putative benefits on patient outcomes from better health services overseas may be mediated by social support. There is a need to understand and explore how social support influences cancer care in the Caribbean. To this date, no qualitative investigation has been conducted comparing social support systems of Caribbean cancer survivors treated overseas with those treated on-island. Preliminary data showed that Saint Lucian survivors depended highly on support from family for a positive cancer experience and may be particularly vulnerable during overseas travel when they are forced to separate from their family [33].

### Cancer care gaps in the SIDS of the OECS

The SIDS that make up the Organization of Eastern Caribbean States (OECS), are particularly susceptible to system-related cancer mortality [34] because of the dearth of radiotherapy facilities in these islands and under-developed health policies for cancer care [35]. The current referral system for overseas care in the OECS may expose patients to unnecessarily long waiting times [36]. However, in the absence of adequate policy, travel restrictions resulting from external events (such as natural disasters and pandemics) and suboptimal processes across the care continuum may continue to contribute to higher cancer mortality. Data on the cancer burden in the OECS region is limited [2, 37] but based on 2022 data from GLOBOCAN for Saint Lucia, we know that the most common cancers are Prostate (93.9), Breast (51.7), Cervical (15.7), and Colon (12.3) [2]. The OECS nationals residing in the islands of Antigua, Dominica, Grenada, Saint Kitts, Saint Lucia and Saint Vincent (∼ 600 000 inhabitants) belong to the same economic union, and have a common political agenda regarding cancer control [38]. Chemotherapy and other pharmaceuticals for these six islands are procured by the OECS commission. On one hand, all these islands have similar capacity in terms of cancer diagnosis, general surgery and medical oncology [1]. On the other hand, there are a few differences in terms of health services offered. e.g. Antigua had radiotherapy (2016-2023) and the oncology units in Saint Kitts and Saint Vincent were recently set up (<10 years) compared to the others. Also, preferences for travel destinations for oncology care are often linked to geographical and socio-cultural proximity. A multicentre approach for epidemiological studies within the OECS will not only ensure greater generalizability and capacity for the implementation of new regional cancer control policies within the OECS but it will help overcome sample size limitations inherent to SIDS.

In order to improve mortality rates and patient outcomes, policymakers need to determine priority areas for better cancer care, and decide between improving access to cancer services from overseas, and developing infrastructure and services on-island. These crucial decisions should be based on scientific evidence examining itineraries for medical travel, characteristics of travellers and the services being accessed overseas. However, overseas travel, in regards to cancer care, remains largely understudied in SIDS [1, 3, 39], and vital epidemiological data on the impact on patient outcomes are lacking.

Recent review articles have presented the treatment landscape of Caribbean SIDS [1, 3, 35]. One review emphasized that models of cancer care from high-income countries (HIC) are not always appropriate for the Caribbean context [1]. Another review reported that approximately hundreds of patients from the of islands Grenada, Dominica and Saint Lucia receive care every year in Martinique, a developed French-Caribbean island [40]. These reviews were pivotal first-works for the Caribbean but like most studies on this topic they were lacking patient-level data (characteristics and lived experience) [1, 3] Our preliminary work showed that in Saint Lucia, a little over half of cancer patients travel for cancer diagnostic services and treatment [5, 33]. Larger studies are still lacking on lived experiences, treatment times, lifestyles and characteristics of Caribbean patients seeking care overseas.

## METHODS/DESIGN

### Research aims & hypotheses

CaSIDEC is an acronym for “Cancer in SIDS of the Eastern Caribbean”. The study’s overarching goal is to investigate the association with overseas travel for care and known determinants of patient outcomes using mixed-methods in Caribbean SIDS, and to contribute towards building research capacity in SIDS. Our primary hypothesis is that care overseas is associated with longer treatment times, less social support and healthier lifestyles post-treatment. We will pursue the following specific aims:

Aim 1: Establish a cohort of 900 cancer survivors in six Caribbean states [face-to-face surveys]:

(a) Investigate treatment delays associated with overseas travel for care among cancer survivors.
(b) Investigate lifestyle factors post-treatment between cancer survivors treated overseas vs. on-island.

Aim 2: Investigate the association between overseas travel for care and social support systems of cancer survivors and caregivers [qualitative interviews & focus groups].

We hypothesize that:

H1a: There are significantly longer treatment delays among persons who get treated overseas compared to those treated on-island.
H1b: Survivors who seek care overseas have lifestyles (i.e. diet and physical activity, substance use) that are more favourable to cancer survival compared to those who seek care on-island.
H2: Compared to survivors who receive cancer care on-island, those who go overseas suffer from social isolation and/or have less social support.

### Overall CaSIDEC study

We will establish a cohort of 900 cancer survivors residing in the islands of Antigua, Dominica, Grenada, Saint Kitts, Saint Lucia and Saint Vincent (Figure 1). Eligible participants will be cancer survivors ≥18 y, able to communicate in English or local dialect (without cognitive impairment), with a cancer diagnosis (any cancer site, histology and diagnosis year) and having accessed health services in their island of residence due to cancer. The proposed study builds on the DCAP study, a pilot that AA conducted in Saint Lucia (*n*=50) (Table 1) [5]. The details on the study design and data from the DCAP study are described in detail elsewhere [5]. Cancer survivors recruited in the pilot study gave their informed written consent before participation. The questionnaire that was developed for the pilot study [5] will be used for Aim 1a. Variable definitions will follow the “*Model of Pathways To Treatment*” [41, 42]. We will study the role of social support in cancer care in Caribbean SIDS by using focus groups and one-on-one interviews of informal caregivers and patients (5 y from diagnosis). Participation will include written authorization to access patient’s data from medical records in health care institutions.

**Figure 1:**
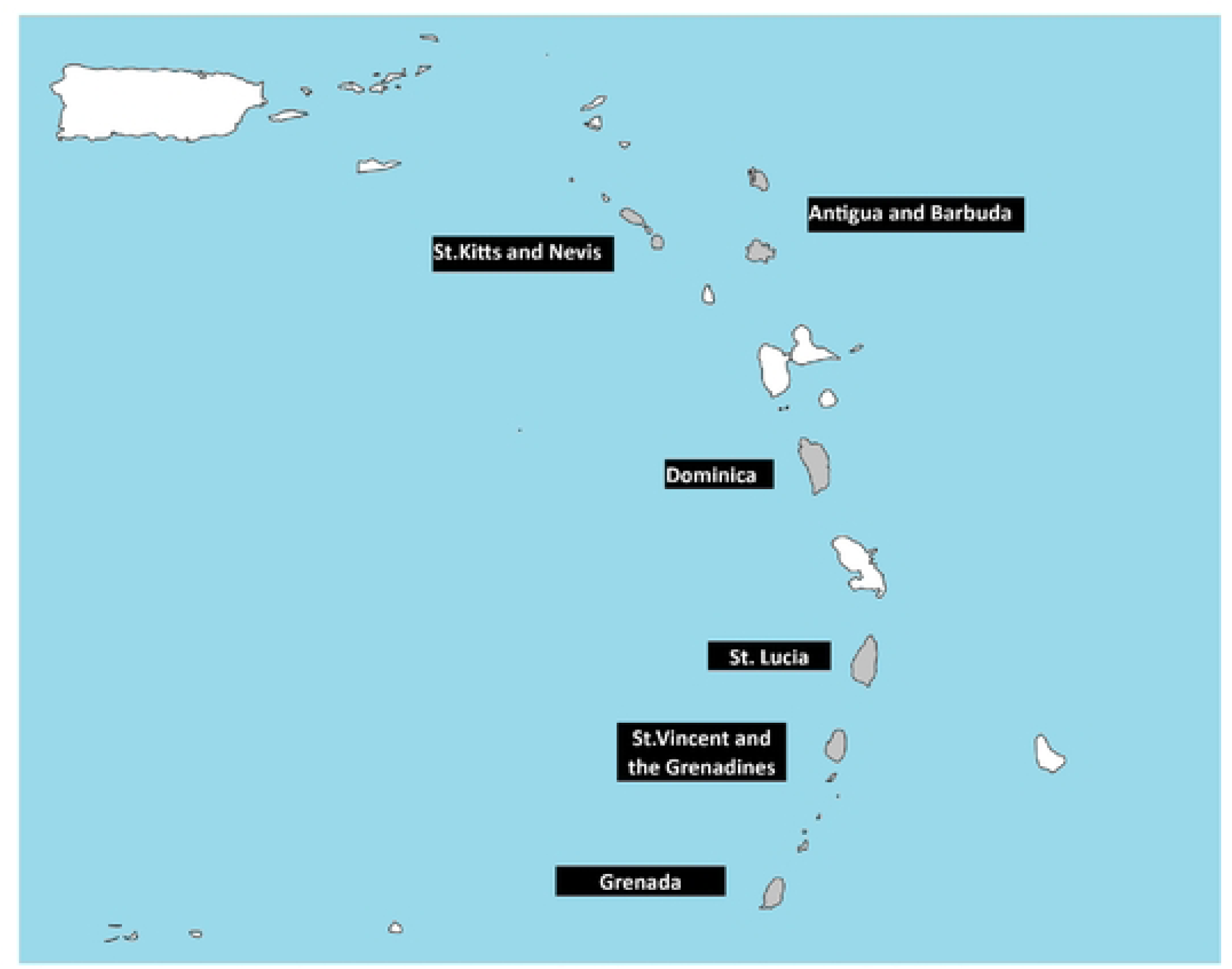
The six island states in the Eastern Caribbean covered by the CaSIDEC study Antigua & Barbuda, Dominica, Grenada, Saint Kitts & Nevis, Saint Lucia, and Saint Vincent & the Grenadines

**Table 1:**
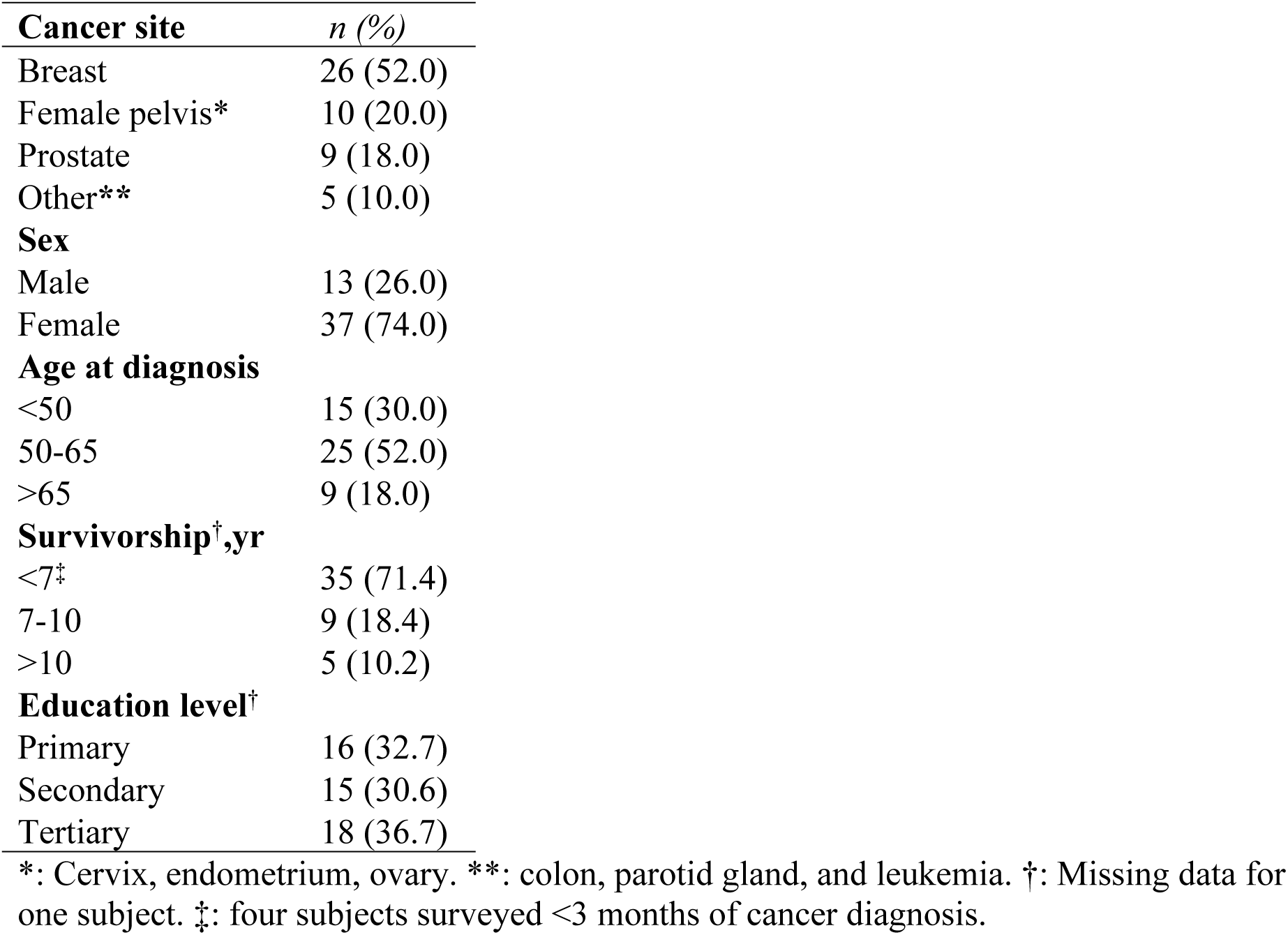
Characteristics of patients included in the DCAP pilot study (n=50)

### Quantitative study on cohort of cancer survivors

#### Study design and sampling

For Aim 1a, time since diagnosis will be limited to 5 years to minimize recall bias (61% of the pilot sample). We will recruit survivors so that the sample characteristics are as representative as possible of all cancer survivors of each island by age, sex, district of residence, income and cancer site. Since there are not any population-based cancer registries in these SIDS, the target distribution for sampling of cancer cases for each island will be developed from data from the World Health Organization (WHO) reports [43], GLOBOCAN, hospital registries (where available) [37], cancer support groups and medical assistance programs. Every six months during data collection, we will generate the proportions of these characteristics in the study sample by island. This routine verification will identify the patient profiles which are over/underrepresented compared to the projected target sample. We will then adjust the recruitment strategy to ensure that our sample conforms to the target distribution.

For Aim 1b, we will conduct a study on a subset of the cohort to investigate the association with overseas travel for care and lifestyle factors post-treatment. We will investigate lifestyle factors within 3 months of ending of active treatment. Participants who finished treatment after 3 months will have two measures for lifestyle: (1) retrospectively at 3 months post-treatment and (2) time of the survey. Eligible participants will be a subset of patients from Aim 1a who already finished their initial active treatment at least 3 months before the time of the survey.

#### Power calculation

A sample of 550 survivors (61% of a cohort of 900) will be sufficient to detect medium differences (β=0.39) in time intervals between persons seeking care overseas and seeking care on-island with a 2-sided α risk of 5% and a statistical power of 90% assuming 10 covariates in the model. Based on pilot data, 67% of patient completed treatment. Therefore, 603 are expected for the sub-sample (Aim 1b). The sample will be sufficiently powered to detect moderate effect sizes (Proportionality-ratio=1.7, 80% power and two-sided 5% alpha risk) using a multivariate model (*G*power software, v3.1.9.4*).

#### Recruitment

Sites for recruitment will be cancer support groups, local hospitals and private oncology clinics. Local clinicians will be trained to actively recruit eligible patients from their practice. Recruitment will be further advertised through social media, public broadcasting media and through the large mailing lists from our cancer support groups. We will also leverage the local medical fraternity organizations, and the Ministries of health to assist in advertising participant recruitment. The cancer support groups will identify eligible participants among their group membership. Where possible, we will recruit cancer survivors (including non-members) during cancer advocacy activities organized by the cancer support groups (health fairs etc.). Patients at health care establishments will be recruited during opportunistic cancer navigation assistance by a representative from a support group. The local oncologists will also invite eligible patients to participate during routine care at health care establishments. Snowball sampling will be used during surveys to identify prospective participants [44].

#### Data collection

Once consented, eligible participants will be contacted by a trained interviewer for a face-to-face survey. Face-to-face is preferred as this reduces issues related to the interpretation of different terms and time intervals commonly associated with self-completion questionnaires in similar research [42]. The pilot survey demonstrated good completeness during the pilot survey. We had less than 10% missing data for the details for help-seeking, medical consultations and treatment. We will ascertain sociodemographics such as education level, private medical insurance, income, hot water at home, employment/education, and clinical characteristics, such as cancer stage at diagnosis, and comorbidities.

We will also collect detailed information from all consultations with health care providers (HCP), from first presentation to initiation of active treatment, including clinical investigations for cancer, specialty of the HCP, location of consultation, scheduling and date of consultation, symptoms, tests and treatments prescribed, referrals, scheduling of review appointments, and suggestions for improving the described management. For each diagnostic test, date, lab name and location, turnaround time, and the finding (whether a test revealed suspicion of cancer) will be ascertained. All treatment modalities will be recorded, including natural/alternative remedies. For each modality, information from the first and last time it was administered (type, date, specialty of physician, location, and country) will be recorded. We will collect information on psycho-social support from loved one and providers (including the Duke-UNC Functional Social Support Questionnaire [45]), and supportive care services accessed.

Funding modalities, fundraising experience and the associated psychological burden will be ascertained. We will ascertain access to palliative care, management of pain and late treatment effects (e.g. lymphedema). Modalities for post-treatment follow-up such as provider specialty, frequency of visits, tests, and health promotion will be collected during the interview. Participants’ personal appraisal of their experiences for major events will be ascertained throughout the interview using a *Likert scale* and open-ended questions [46]. For every country visited for diagnosis and/or treatment, we will record the services accessed, and motives for the choice of country (personal/HCP preference vs. lack of infrastructure).

Patients will complete the NCI-Food Frequency Questionnaire (FFQ) [47]. Other traditional/alternative medicines consumed will be ascertained. Special attention will be given to tisanes from local plants such as soursop (*Annona muricata L*.) and papaya (*Carica papaya L.*) which are consumed by cancer survivors from the Caribbean [5] and have anti-cancer properties [48, 49]. Moderate– to vigorous–intensity Physical activity (PA) will be self-reported by the validated SQUASH questionnaire (Short QUestionnaire to ASsess Health-enhancing physical activity) [50]. Body mass index (BMI) at the time of the interview will be ascertain using a scale and stadiometer. Waist circumference will be measured with a measuring tape by a trained field investigator. Self-report will be used for retrospective measures. *Substance use:* We will assess tobacco status and alcohol drinking.

We will collect clinical data from the patients’ medical records from the various health care establishments (private and public), and medical laboratories on-island. Data from medical records will help prevent issues related to data completeness and misclassification bias from self-report. We will abstract specifically, clinicopathological characteristics of the cancer (anatomical stie, TNM staging, and histology), date of lab results and cancer treatments.

#### Statistical analysis

We will use multivariable linear regressions to estimate mean time intervals, as a function of overseas travel for treatment. Overseas travel will be treated as a binary variable (Travel for treatment vs. ref=treatment exclusively on-island). Medical records data will be used to calculate the time from diagnosis (biopsy date) to treatment (first treatment date). To better interpret the potential clinical impact of our results, we will also operationalize the time to treatment variable as the number of days from recommended benchmarks based on scientific evidence (e.g. UK’s National Health Service). We will assess the presence of mediation by socioeconomic status and social support by estimating the direct and indirect effects proposed by Valeri and VanderWeele [51].

We will calculate overall lifestyle scores based on cancer prevention recommendations from the World Cancer Research Fund/American Institute for Cancer Research (WCRF/AICR) [52]. *DHQ III nutrient database* will be used to derive the nutrient values from the FFQ [47]. Quantitative cutoff points will be used to determine adherence to the different components from the guidelines. The quantitative cutoff points correspond mostly to values which significantly influence risk of cancer, non-communicable diseases and all-cause mortality [52, 53]. We will use linear regression models to estimate the association between country of treatment (overseas vs. on-island) and healthy lifestyle. For these models, a 1-SD increase in each lifestyle score will be calculated to allow comparability between the scores. To achieve more clinically relevant interpretations, the scores will be dichotomized based on cutoff points determined by recommendations based on reducing all-cause mortality among cancer survivors [52]. The association with this variable will be estimated from proportionality-ratios (PR) and their 95% confidence intervals (CI) using multivariable log-binomial regressions.

All models will include age at diagnosis, cancer site, stage at diagnosis, gender, country of residence, socioeconomic status and years since diagnosis (minimally adjusted). Models for Aim 1b will be further adjusted for treatment type, comorbidities, health promotion accessed. We will also consider using multiple imputations to manage missing data. we will also use a directed acyclic graphs (DAGs) to help identify potential confounders [51]. An *a priori* alpha level of 0.05 will be used to determine statistical significance. We intend to analyse all participants together to maximize statistical power. However, we acknowledge that treatment waiting times and lifestyles may vary according to certain innate characteristics such as treatment protocols specific to cancer sites and country of residence, cultural differences, treatment location and patients’ gender.

Sensitivity analyses will be performed to assess the impact of these potential variations on results and the study’s robustness. We will examine effect modifications and perform subgroup analyses by covariates listed in minimally adjusted model. To minimise recall bias, we will systematically conduct analyses restricted to patients who were diagnosed more recently (1a: ≤12 months & 1b:≤24 months). We will introduce the country of residence as a covariable or as a random effect in the regression model. Where possible, we will also perform analyses by country to highlight country-specific features.

### Qualitative study on social support

#### Design

*One-on-one interviews* will be held with 24 cancer survivors across the six islands. We will approach survivors according to medical travel status. We will interview survivors treated exclusively on-island (*n*=12), treated partially overseas (*n*=6) and survivors treated entirely overseas (*n*=6). *Focus groups* will capture the synergistic relationships among participants from the six islands, and enable immediate cross-country comparisons. The caregivers of patients will also be invited to participate and interviewed separately from the survivors. Caregivers will be recruited using means similar to patients: i.e. advertisement and leveraging the networks of support groups. Six focus groups consisting of 6–9 survivors/caregivers will be conducted (*n*=∼54). Members of the focus group may be distinct from interviews. Members will have the same medical travel status. The total sample sizes are comparable to other similar qualitative studies [39, 54, 55]. We will purposefully identify [56] cancer survivors and caregivers of survivors who satisfy these criteria: (1) are receiving or already finished their active treatment at the time of the study, (2) received a treatment requiring the intervention of a medical professional.

#### Data collection

*One-on-one interviews* will be initiated by an introductory question “describe the support you received during the cancer journey”, then guided by questions and prompts “who”, “what was the relationship to you? such as a family, friend, neighbours" and “when”. Participants will be encouraged to focus on aspects of the day-to-day logistics and travel arrangements, experience living overseas. All interviews will be audio recorded using Zoom. Interviews will be transcribed by investigators using the live transcription feature in Zoom [57]. Qualitative interviews will be organized and scheduled six-months following start of the survey from Aim 1. A member of our team with qualitative research experience will moderate the *focus group* in English using a semi-structured, open-ended focus group interview guide [58], while a second co-investigator takes notes. Six focus groups in total (3 for survivors and 3 for caregivers). Each country will contribute 3-4 participants max per focus group (6-9 persons). Discussions will be held at community centres, cancer support group building or virtually using a suitable video-conferencing tool (e.g. MS Teams, Zoom). Our focus group questions will follow the model proposed by Ruff et al. [59], using open-ended questions that will include introductory questions, key questions, and a concluding question. Introductory questions (e.g. “Can you talk about the process leading up to your cancer treatment?”), will encourage participants to respond from their own experiences. Key questions will focus on administrative procedures, travel logistics and lived experience in the country(ies) where they received treatment, with questions such as, “What was it like leaving your home country for treatment?”, “How did you feel about leaving your family and loved one back home?”, “What were some of changes you experienced when you arrived in the destination country for treatment (e.g. change in climate)?”. We will ask concluding questions to ensure that complete information had been obtained. At the end, we will ask, “Is there anything else you would like to say about being in a foreign country for cancer treatment?”.

#### Data analysis and management

Transcripts will be initially analysed through the grounded theory approach, a data-driven, open coding process [60]. This will be supplemented with subsequent concept-driven coding [61]. Two of our authors independently will analyse and code the same sample of responses. Interview responses will be blinded for the other variables (age, **gender**, cancer site etc.) to prevent them from influencing the research findings. Furthermore, the decisional criteria that emerged in the previous focus groups will be reported to the subsequent ones at the end of discussions, with the intention of introducing new insights and data [62]. Five years after the studies completion, data and underlying reported findings will be shared with the African-Caribbean Consortium for access to all its members.

### Ethics

This protocol was approved by the McGill University’s Institutional Review Board (ID: A10-M79-24A). Ethics approval was also granted by the Antigua and Barbuda Institutional Review Board (ID: AL-01/102024-ANUIRB), Ethics committees of the Commonwealth of Dominica (ID: N/A), St.George’s University IRB (Grenada, ID: 23005), The research ethics committee of the Medical and Dental Council (Saint Lucia, ID: N/A), The Ministry of Health Ethics Review Committee of St.Kitts (MOH-ERC-2024-09-072). The study will be conducted in compliance with Good Clinical Practice Procedures, the principles of the Declaration of Helsinki. Participants who meet the eligibility criteria who wish to take part in the study after they have been informed about its procedures, they will be asked to sign an informed consent form.

### Timing

The project is currently in the planning and development phases. Completion of the logistics and procedures to launch participant recruitment across these sites is projected for the last quarter of 2025. We estimate that participant recruitment and data collection will be completed on the 31^st^ of October 2028. The final results of the CaSIDEC study are expected within two years of data collection.

## DISCUSSION

The proposed research is innovative because its addresses a potential determinant of cancer mortality specific to a much-understudied population [11, 63]. The status quo as it pertains to research on overseas travel for cancer care is studies focusing on patients from large LMICs. Furthermore, these studies draw data largely from self-reports at the provider level, clinical registries and government sponsorship programs. Thus, these studies do not address patient outcomes, their views or quality of care. Only recently has patient-centred outcomes been investigated among patients from SIDS [5, 33, 39].

We will investigate for the first time, overseas travel for cancer care in multiple Caribbean SIDS. We will focus on major knowledge gaps; notably, care pathways, social support and patient-centred outcomes. The data from this proposal are expected to drive the development of strategies to optimize the overseas referral process taking into account the psychological well-being of patients and caregivers. Shorter delays for treatment, funding, travel approval and arrangement along with additional social support at key moments will ultimately help prevent cancer deaths in SIDS.

The main limitation of this study will be that we are unable to use random sampling or recruit solely incident cases, which may introduce selection bias toward less severe, long-term survivors. There are no population-based cancer registries in the OECS to facilitate sampling. To mitigate this, we will systematically recruit newly diagnosed cancer patients via oncology clinics and NGOs. While recall bias is a limitation when including prevalent cases, sensitivity analyses restricted to those diagnosed within 12 months will reduce this risk. Despite these constraints, with an estimated 3,000+ prevalent cases across the six islands (GLOBOCAN, 2022), our target of enrolling 900 participants over three years remains feasible [2].

### Knowledge translation and future interventions

Our work will produce recommendations to fill performance gaps in the systems for cancer care in the OECS that will improve patients’ lives at multiple phases. It is an important first step to developing data resources for high-quality research in understudied SIDS.

We plan to inform change in practice and policy. We will implement a robust dissemination and knowledge transfer strategy rooted in community organizing principles. One of the outputs to be considered will be OECS-wide treatment guidelines recommending destinations based on the patient-centred outcomes and waiting times reported from our study. CaSIDEC’s Steering Committee (Figure 2) will work in concert with policymakers to ensure actionable improvements in cancer care. Our strategy includes using the Caribbean Cancer Portal, to disseminate our research findings, share best practices, and provide real-time updates to survivors, healthcare professionals, and policymakers. We will foster interdisciplinary dialogue on critical issues like treatment delays, diet, physical activity and social support systems during/after cancer. via a series of conferences, seminars, webinars, and workshops. Our steering committee will meet regularly to discuss the advancement in science with local stakeholders. We have also integrated knowledge user and health policy experts in Fiji to further strengthen our capacity for knowledge translation. Their involvement will encourage cross-pollination with practices in other small islands context and help emerge innovative concepts.

**Figure 2.**
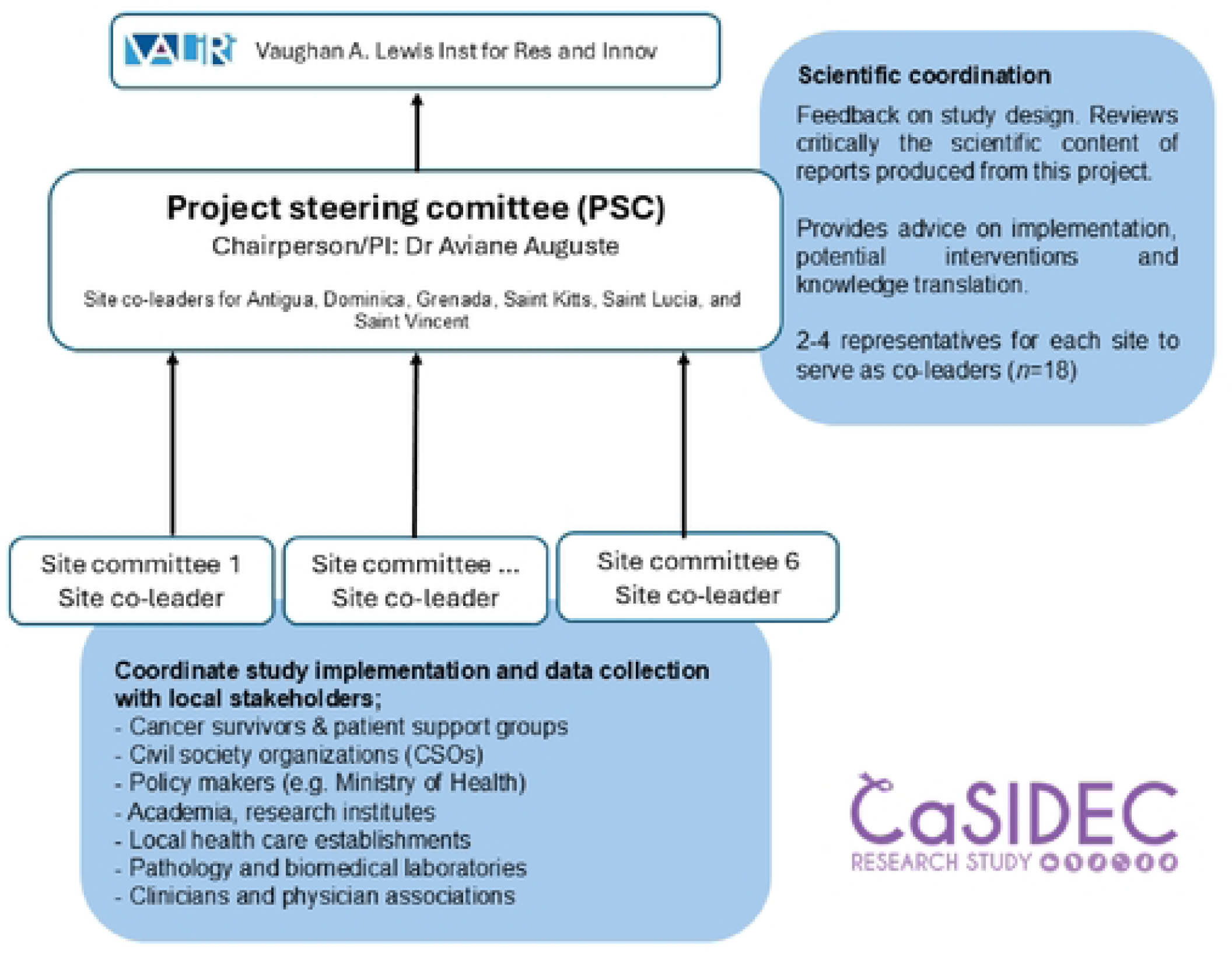
Project governance for the CaSIDEC study

## Data Availability

All relevant data are within the manuscript

## DECLARATIONS

### Ethics approval and consent to participate

This protocol was approved by the McGill University’s Institutional Review Board (ID: A10-M79-24A). Ethics approval was also granted by the Antigua and Barbuda Institutional Review Board (ID: AL-01/102024-ANUIRB), Ethics committees of the Commonwealth of Dominica (ID: N/A, 2025), St.George’s University IRB (Grenada, ID: 23005, 2023), The research ethics committee of the Medical and Dental Council (Saint Lucia, ID: N/A, 2023), The Ministry of Health Ethics Review Committee of St.Kitts (ID: MOH-ERC-2024-09-072). Participants who meet the eligibility criteria who wish to take part in the study after they have been informed about its procedures, they will be asked to sign an informed consent form. The data presented in the pilot were derived from the DCAP study, were approved by the Ethics Committee of the Medical and Dental Council (Saint Lucia, ID: N/A, 2019).

### Data availability

Five years after the studies completion, data and underlying reported findings will be shared with the African-Caribbean Consortium for access to all its members.

### Consent for publication

Not applicable

### Author contributions

AA conceived and designed the original study. AA, EB, CH, JQ, HY, YA, LL, NS, SN, LT, JMD, MN, SDW, OG, DP, AD, JMR, TW, TB contributed to the study design and data collection tools. AA, EB, HY, YA, LL, SN, LT, MN, SDW, OG, DP, AD, JMR, TW, TB contributes to the data collection. AA leads the funding acquisition for the study. AA, LL, TB participate in the coordination and logistics of the study. AA, HY, YA, SN, LT, MN, SDW, OG, DP, AD, JMR, TW, TB coordinate the recruitment of cancer patients in the respective countries. AA drafted the manuscript and prepared Figures 1 and 2. All authors agreed with the final version of the manuscript and critically revised the manuscript.

## ACKNOWLEDGEMENTS

We thank Dr Camille Ragin, JoAnn Oliver from the African-Caribbean Cancer Consortium (AC3), and Dr Erin Strumpf (McGill University) for their mentorship to the early career investigators on the CaSIDEC study team. We extend our sincere gratitude to Dr Roxanne Brizan-St. Martin (Programme Director, Health and Social Inclusion, OECS Commission) for her steadfast institutional support and regional guidance in communicating with policymakers for knowledge translation. We gratefully acknowledge Dr Carole Naidu, Amelia Hyatt, Belinda Chan, Etivina Lovo, and Chris Bates from Fiji for their invaluable contributions to mutual learning and cross-regional knowledge translation between the Pacific and Caribbean, which have enriched the dissemination plan of this research.

To the Ministry officials, policymakers and their staff. we are thankful their facilitation of local research approvals: Mrs. Ena Dalso Henry, Collin O’Keefe, Dr. Shawn Charles, Dr Laura Esprit, Hon. Cassanni Laville, Linel Augustine, Isha George, Dr Hazel Laws, Sheneil Isles, Dr Sherry Ephraim, Dr Glensford Joseph, Dr Michelle Francois, Nurse Yolanda Alcindor, Shanna Philbert-Cyr, Dr. Damian Greaves, Shamanti Labban, Mr. Cuthbert Knights, Dr. Simone Keizer-Beache, Hon Dr Ingrid Buffonge, Guy-Albert Rufin-Duhamel, Fatiha Nehal, and Dr Miguelle Marous. We appreciate the partnership of cancer support group members: Erma Lloyd, Davis Letang, Delia Cuffy-Weekes, Debbie Martin Jacques, Don Coriette, Alecia Bonadie, Patricia Farell and Mrs. Pamela Hendrickson—for their community engagement, survivor advocacy, and commitment to patient-centered research.

Our thanks to the dedicated field investigators and interviewers for volunteering to do the field work for this study: Icilma Wilkin, Ronasha Williams, Cecile James, Wonderful Okunade, Gifta Jongue, Margueritte Annette Jn Charles, and Euphemia Edmund. We also recognize the continued contributions of our colleagues at VALIRI for their leadership, coordination, and operational support in Saint Lucia: Ing. Verne Emmanuel, Dr Merle St. Clair-Auguste, and Dr Winston Phulgence. We are grateful to Omar Combie and Nathalie Jolie-Fanis from the Marketing Unit at Sir Arthur Lewis Community College for their expertise in communications and public outreach.

Finally, we thank the medical and paramedical professionals: Dr. Tamara Remy, Rudy Avril, Dr Asha Martin, Nurse Darrel John, Dr Wayne Felicien, Dr Stephen King, Irenoeus Descartes, Dr Naveen Urs, Dr Dawit Kabiye, Dr Diana Callendar, Dr Humberto Gamez for their insights into the realities of cancer care in the Eastern Caribbean.

## CaSIDEC study group: Project steering committee

Eunetta Bird, Celine Heskey, Jaqui Quinn, Hanybal Yazigi (Antigua), Yvonne Alexander, Lyndelle LeBruin, Nick Shillingford (Dominica), Sonia Nixon, Lindonne Telesford (Grenada), Janielle P. Maynard, Marcus Natta, Steve D. Whittaker (Saint Kitts and Nevis), Owen Gabriel, Dorothy Phillip (Saint Lucia), Ariane Duncan, Jozelle Miller, Tami Williams (Saint Vincent and the Grenadines). ***Vaughan A. Lewis Institute for Research and Innovation (VALIRI):*** Tricia Black

## Authors’ information

AA is an Epidemiologist from Saint Lucia. He is currently an Assistant Professor at McGill University as well as a researcher and member of the leadership of the Vaughan A. Lewis Institute for Research and Innovation (VALIRI), a multi-disciplinary research institute established by the Sir Arthur Lewis Community College in Saint Lucia. All members of the CaSIDEC Study Group, including the project’s steering committee, are Afro-Caribbean nationals originally from the OECS Member States included in this study protocol. This composition reflects an intentional commitment to conducting ethically grounded global health research that is locally led and regionally.

## ABBREVIATIONS

AICR: American Institute for Cancer Research
BMI: Body mass index
CaSIDEC: Cancer in small island developing states of the Eastern Caribbean
CI: Confidence intervals
DAG: Directed acyclic graphs
FFQ: Food frequency questionnaire
GIRC: Geographically isolated and resource-constrained
HCP: Health care providers
HIC: High-income countries
LMIC: Low- and middle-income country
OECS: Organization of Eastern Caribbean States
PA: Physical activity
PR: Proportionality-ratios
SIDS: Small Island Developing States
SQUASH: Short QUestionnaire to ASsess Health-enhancing physical activity
WCRF: World Cancer Research Fund
WHO: World Health Organization

